# Infection fatality risk for SARS-CoV-2: a nationwide seroepidemiological study in the non-institutionalized population of Spain

**DOI:** 10.1101/2020.08.06.20169722

**Authors:** Roberto Pastor-Barriuso, Beatriz Pérez-Gómez, Miguel A Hernán, Mayte Pérez-Olmeda, Raquel Yotti, Jesús Oteo-Iglesias, Jose L Sanmartín, Inmaculada León-Gómez, Aurora Fernández-García, Pablo Fernández-Navarro, Israel Cruz, Mariano Martín, Concha Delgado-Sanz, Nerea Fernández de Larrea, Jose León Paniagua, Juan F Muñoz-Montalvo, Faustino Blanco, Amparo Larrauri, Marina Pollán, on behalf of the ENE-COVID Study Group

**Affiliations:** National Centre for Epidemiology, Institute of Health Carlos III, Madrid, Spain; Consortium for Biomedical Research in Epidemiology and Public Health (CIBERESP), Institute of Health Carlos III, Madrid, Spain; Departments of Epidemiology and Biostatistics, Harvard T.H. Chan School of Public Health; Harvard-MIT Division of Health Sciences and Technology, Boston, MA, USA; National Centre for Microbiology, Institute of Health Carlos III, Madrid, Spain; Institute of Health Carlos III, Madrid, Spain; Spanish Network for Research in Infectious Diseases (REIPI), Institute of Health Carlos III, Madrid, Spain; Ministry of Health, Madrid, Spain; National School of Public Health, Institute of Health Carlos III, Madrid, Spain

## Abstract

**Objective:** To estimate the range of the age- and sex-specific infection fatality risk (IFR) for severe acute respiratory syndrome coronavirus 2 (SARS-CoV-2) based on confirmed coronavirus disease 2019 (COVID-19) deaths and excess all-cause deaths.

**Design:** Nationwide population-based seroepidemiological study combined with two national surveillance systems.

**Setting and participants:** Non-institutionalized Spanish population of all ages.

**Main outcome measures:** The range of IFR was calculated as the observed number of COVID-19 deaths and excess deaths divided by the estimated number of SARS-CoV-2 infections in the non-institutionalized Spanish population. Laboratory-confirmed COVID-19 deaths were obtained from the National Epidemiological Surveillance Network (RENAVE) and excess all-cause deaths from the Monitoring Mortality System (MoMo) up to July 15, 2020. SARS-CoV-2 infections were derived from the estimated seroprevalence by a chemiluminiscent microparticle immunoassay for IgG antibodies in 61,092 participants in the ENE-COVID nationwide serosurvey between April 27 and June 22, 2020.

**Results:** The overall IFR (95% confidence interval) was 0.8% (0.8% to 0.9%) for confirmed COVID-19 deaths and 1.1% (1.0% to 1.2%) for excess deaths. The IFR ranged between 1.1% (1.0% to 1.2%) and 1.4% (1.3% to 1.5%) in men and between 0.6% (0.5% to 0.6%) and 0.8% (0.7% to 0.8%) in women. The IFR increased sharply after age 50, ranging between 11.6% (8.1% to 16.5%) and 16.4% (11.4% to 23.2%) in men ≥80 years and between 4.6% (3.4% to 6.3%) and 6.5% (4.7% to 8.8%) in women ≥80 years.

**Conclusion:** The sharp increase in SARS-CoV-2 IFR after age 50 was more marked in men than in women. Fatality from COVID-19 is substantially greater than that reported for other common respiratory diseases such as seasonal influenza.

**WHAT IS ALREADY KNOWN ON THIS TOPIC:** Infection fatality risk (IFR) for SARS-CoV-2 is a key indicator for policy decision making, but its magnitude remains under debate. Case fatality risk, which accounts for deaths among confirmed COVID-19 cases, overestimates SARS-CoV-2 fatality as it excludes a large proportion of asymptomatic and mild-symptomatic infections. Population-based seroepidemiological studies are a valuable tool to properly estimate the number of infected individuals, regardless of symptoms. Also, because ascertainment of deaths due to COVID-19 is often incomplete, the calculation of the IFR should be complemented with data on excess all-cause mortality. In addition, data on age- and sex-specific IFR are scarce, even though age and sex are well known modifiers of the clinical evolution of COVID-19.

**WHAT THIS STUDY ADDS:** Using the ENE-COVID nationwide serosurvey and two national surveillance systems in Spain, this study provides a range of age- and sex-specific IFR estimates for SARS-CoV-2 based on laboratory-confirmed COVID-19 deaths and excess all-cause deaths. The risk of death was very low among infected individuals younger than 50 years, but it increased sharply with age, particularly among men. In the oldest age group (≥80 years), it was estimated that 12% to 16% of infected men and 5% to 6% of infected women died during the first epidemic wave.

## INTRODUCTION

The infection fatality risk (IFR)—the proportion of infected individuals who die from the infection—is a key indicator to design public health policies to control infectious diseases. Because the magnitude of the IFR for severe acute respiratory syndrome coronavirus 2 (SARS-CoV-2) remains under debate,^1,2^ lockdowns and other extreme forms of social distancing have been questioned as appropriate responses to the coronavirus disease 2019 (COVID-19) pandemic.

An accurate estimation of the IFR of SARS-CoV-2 is difficult. Even if all symptomatic infections were diagnosed, something that so far has not occurred in most countries, asymptomatic infections cannot be clinically identified. Therefore, estimating the IFR needs to rely on population-based serosurveys that provide an estimate of the proportion of individuals that has been infected, regardless of symptoms.^3^ Also, because ascertainment of deaths attributable to COVID-19 is often incomplete, the calculation of the IFR can be complemented with data on excess mortality.

A recent unpublished review of 24 serological reports^4^, several of them also unpublished, estimated an overall IFR of 0.68% (95% confidence interval 0.53% to 0.83%). However, the methodological quality of many of these studies was questionable, with some exceptions^5^. IFR estimates were mostly based only on surveillance-registered deaths, and there was a very high between-study heterogeneity, with estimates ranging from 0.16% to 1.60%. Also, because the IFR for SARS-CoV-2 is expected to increase with age and may differ by sex, overall crude IFR estimates cannot be directly compared between populations with different age and sex structure (e.g., China and Western Europe). Accurate and reliable age- and sex-specific estimates of IFR are urgently needed.

Here, we report overall and age- and sex-specific IFR estimates for SARS-CoV-2 from ENE-COVID, a large nationally representative serosurvey in the non-institutionalized Spanish population.

## METHODS

### Estimation of the number of SARS-CoV-2 infections

We calculated the prevalence of IgG antibodies against SARS-CoV-2 in the non-institutionalized Spanish population using data from ENE-COVID, a nationwide population-based serosurvey whose design has been described elsewhere.^6^ Briefly, 1,500 census tracts, and up to 24 households within each tract, were randomly selected using a two-stage sampling stratified by province and municipality size. All residents of the 35,883 selected households were invited to participate in the study, which was carried out between April 27 and June 22, 2020 in three two-week rounds, with a one-week break between rounds. Epidemiologic questionnaires and serology tests were administered to 68,292 individuals who participated in at least one round.^7^ The study used two immunoassays to detect IgG antibodies: a point-of-care test (Orient Gene Biotech COVID-19 IgG/IgM Rapid test Cassette), and a chemiluminiscent microparticle immunoassay (CMIA) that required venipuncture (SARS-CoV-2 IgG for use with ARCHITECT; Abbott Laboratories, Abbott Park, IL, USA; reference 06R8620), with better performance characteristics (see supplementary methods and supplementary figure 1 for a summary of reported sensitivity and specificity estimates of the CMIA test).^6^

We calculated the seroprevalence, overall and in strata defined by age and sex, as the proportion of participants who had detectable IgG antibodies against SARS-CoV-2 in any round by the CMIA test (61,092 participants had a valid CMIA result). To account for the different sampling selection probabilities by province and to adjust for non-response to the CMIA test based on sex, age, and census tract average income, we assigned sampling weights to each study participant. Design-based standard errors for seroprevalences were computed taking into account the stratification by province and municipality size group and the clustering of seropositivity by household and census tract.^6^

In sensitivity analyses, we corrected the SARS-CoV-2 seroprevalence estimates for the CMIA’s sensitivity and specificity, which were estimated to be 90.6% (95% confidence interval 88.1% to 92.6%) and 99.3% (99.0% to 99.5%), respectively, from a meta-analysis of 23 diagnostic accuracy studies (see supplementary methods and supplementary figure 1 for details).

We calculated the number of seropositive persons in Spain by multiplying the age- and sex-specific prevalences of IgG antibodies times the size of the corresponding non-institutionalized Spanish population groups as of July 15, 2020.^8^

### Estimation of the number of deaths due to COVID-19

Given the practical difficulties in reporting and adjudicating deaths from COVID-19 during the epidemic, we estimated the IFR separately using confirmed COVID-19 deaths and excess all-cause deaths.^9^ The two sources of information were the Spanish National Epidemiological Surveillance Network (RENAVE) and the Monitoring Mortality System (MoMo).

RENAVE^10,11^ provided individual data on the 29,137 laboratory-confirmed COVID-19 deaths registered in Spain up to July 15, 2020. There were 249 deaths (0.9%) with missing demographic data, which were distributed according to the observed sex and age group distribution of all other deaths.

The median interval between onset of symptoms and death in RENAVE data was 12 days (interquartile range 7 to 19 days).

MoMo collects information on deaths from 3,945 municipal civil registries that cover 93% of the Spanish population.^12^ Using a model described elsewhere,^13^ MoMo data are used to quantify excess deaths for a particular period, taking into account the historical series of the last 10 years and incorporating a secular trend and a seasonal component. Between March 1 and July 15, 44,459 excess all-cause deaths were estimated (mainly concentrated between March 13 and May 22).^12^

Neither RENAVE nor MoMo distinguish between institutionalized and non-institutionalized population. It was estimated that 9,909 deaths with confirmed COVID-19 and 19,681 deaths attributed to suspected cases occurred in long-term care facilities, mainly nursing homes, during the same period (supplementary table 1). We subtracted these deaths from those identified by RENAVE and MoMo, respectively, in the population aged 60 years and older (see supplementary methods for details).

### Estimation of infection fatality risks

The IFR is the number of deaths due to COVID-19 divided by the number of individuals with SARS-CoV-2 infection. We obtained separate estimates of the overall IFR using the COVID-19 deaths from RENAVE (lower bound of deaths, due to limited ascertainment in surveillance) and the excess all-cause deaths from MoMo (a possible upper bound because of the inclusion of deaths that may not result from direct or indirect effects of the epidemic). We then repeated the above analyses in each stratum defined by sex and 10-year age group. We calculated 95% confidence intervals based on delta methods that accounted for both the binomial variance in the number of deaths and the estimated design-based variance in the number of infections (see supplementary methods for details). Analyses were carried out using survey commands in Stata, version 16 and survey package in R, version 3.

## RESULTS

According to the ENE-COVID study, the SARS-CoV-2 seroprevalence (95% confidence interval) was 4.9% (4.6% to 5.3%) during the first epidemic wave in Spain, which corresponded to 2.3 million (2.2 to 2.5 million) non-institutionalized individuals with antibodies against SARS-CoV-2 (table 1). Through July 15, 2020, 19,228 laboratory-confirmed COVID-19 deaths and 24,778 excess all-cause deaths were estimated to occur among individuals residing in Spain outside of nursing homes. The distribution by age and sex was similar for both sources of death data: 64% of the COVID-19 deaths and 62% of the excess deaths occurred among men; 79% of confirmed COVID-19 deaths and 83% of excess deaths occurred among individuals aged 70 years or older.

**Table 1.** Infection fatality risk for SARS-CoV-2 in non-institutionalized population by sex and age, ENE-COVID study, April 27–June 22, 2020, Spain.

| Sex, age (years) | Individuals in population, thousands | SARS-CoV-2 seroprevalence*, % (95% CI) | Individuals with SARS-CoV-2 antibodies, thousands (95% CI) | Confirmed COVID-19 deaths | Excess all-cause deaths | Infection fatality risk, % (95% CI) |  |
| --- | --- | --- | --- | --- | --- | --- | --- |
|  |  |  |  |  |  | Based on confirmed COVID-19 deaths | Based on excess all-cause deaths |
| Overall | 46,887.1 | 4.9 (4.6 to 5.3) | 2,306.7 (2,153.6 to 2,470.1) | 19,228 | 24,778 | 0.83 (0.78 to 0.89) | 1.07 (1.00 to 1.15) |
| Men | 23,006.9 | 4.8 (4.4 to 5.2) | 1,106.0 (1,017.6 to 1,201.6) | 12,317 | 15,480 | 1.11 (1.02 to 1.21) | 1.40 (1.29 to 1.52) |
| 0–9 | 2,205.5 | 3.2 (1.9 to 5.4) | 71.7 (42.5 to 119.7) | 3 | 32 | 0.00 (0.00 to 0.01) | 0.04 (0.02 to 0.08) |
| 10–19 | 2,557.9 | 3.7 (2.8 to 4.8) | 93.5 (71.2 to 122.5) | 3 | 0 | 0.00 (0.00 to 0.01) | 0.00 (0.00 to 0.01) |
| 20–29 | 2,479.1 | 5.8 (4.7 to 7.1) | 142.9 (116.2 to 175.3) | 18 | 0 | 0.01 (0.01 to 0.02) | 0.00 (0.00 to 0.01) |
| 30–39 | 2,978.7 | 4.7 (3.8 to 5.7) | 139.7 (114.0 to 170.9) | 48 | 3 | 0.03 (0.02 to 0.05) | 0.00 (0.00 to 0.01) |
| 40–49 | 3,916.7 | 5.3 (4.6 to 6.2) | 209.0 (180.0 to 242.4) | 192 | 168 | 0.09 (0.07 to 0.11) | 0.08 (0.06 to 0.10) |
| 50–59 | 3,493.8 | 5.3 (4.5 to 6.1) | 184.0 (157.8 to 214.3) | 705 | 601 | 0.38 (0.32 to 0.45) | 0.33 (0.27 to 0.39) |
| 60–69 | 2,598.2 | 4.9 (4.1 to 5.9) | 127.2 (105.3 to 153.3) | 1,904 | 2,065 | 1.50 (1.23 to 1.81) | 1.62 (1.34 to 1.97) |
| 70–79 | 1,783.7 | 4.7 (3.7 to 6.0) | 83.7 (65.5 to 106.7) | 4,145 | 5,114 | 4.95 (3.86 to 6.32) | 6.11 (4.77 to 7.79) |
| ≥80 | 993.3 | 4.6 (3.2 to 6.5) | 45.6 (31.8 to 64.9) | 5,299 | 7,497 | 11.6 (8.06 to 16.5) | 16.4 (11.4 to 23.2) |
| Women | 23,880.1 | 5.0 (4.7 to 5.4) | 1,200.5 (1,110.5 to 1,297.4) | 6,911 | 9,298 | 0.58 (0.53 to 0.62) | 0.77 (0.71 to 0.84) |
| 0–9 | 2,078.3 | 4.2 (2.7 to 6.7) | 88.0 (55.1 to 139.0) | 2 | 11 | 0.00 (0.00 to 0.01) | 0.01 (0.01 to 0.03) |
| 10–19 | 2,396.7 | 4.4 (3.4 to 5.6) | 105.1 (81.7 to 134.7) | 3 | 22 | 0.00 (0.00 to 0.01) | 0.02 (0.01 to 0.03) |
| 20–29 | 2,404.1 | 5.7 (4.6 to 7.0) | 137.4 (111.2 to 169.3) | 17 | 10 | 0.01 (0.01 to 0.02) | 0.01 (0.00 to 0.01) |
| 30–39 | 3,012.4 | 5.2 (4.4 to 6.2) | 156.7 (132.0 to 185.8) | 29 | 71 | 0.02 (0.01 to 0.03) | 0.05 (0.03 to 0.06) |
| 40–49 | 3,877.8 | 5.3 (4.6 to 6.2) | 206.8 (177.9 to 240.0) | 103 | 91 | 0.05 (0.04 to 0.06) | 0.04 (0.03 to 0.06) |
| 50–59 | 3,563.5 | 5.2 (4.5 to 6.0) | 184.4 (158.8 to 213.8) | 318 | 369 | 0.17 (0.14 to 0.21) | 0.20 (0.17 to 0.24) |
| 60–69 | 2,803.4 | 5.0 (4.2 to 6.0) | 140.4 (117.2 to 167.9) | 749 | 875 | 0.53 (0.44 to 0.65) | 0.62 (0.51 to 0.75) |
| 70–79 | 2,138.1 | 4.6 (3.7 to 5.8) | 98.9 (79.0 to 123.4) | 1,986 | 2,646 | 2.01 (1.60 to 2.52) | 2.68 (2.13 to 3.35) |
| ≥80 | 1,605.8 | 5.0 (3.7 to 6.8) | 80.2 (58.7 to 108.9) | 3,704 | 5,203 | 4.62 (3.38 to 6.29) | 6.49 (4.74 to 8.82) |

\* Proportion of participants with detectable IgG antibodies against SARS-CoV-2 in any round of the ENE-COVID study by the chemiluminiscent microparticle immunoassay.

Overall, the IFR estimate (95% confidence interval) was 0.83% (0.78% to 0.89%) for confirmed COVID-19 deaths and 1.07% (1.00% to 1.15%) for excess deaths. The corresponding estimates were 1.11% (1.02% to 1.21%) and 1.40% (1.29% to 1.52%) for men, and 0.58% (0.53% to 0.62%) and 0.77% (0.71% to 0.84%) for women (table 1).

The IFR estimate varied greatly with age. It was under 1 per 1000 through age 49, with much lower values in younger age groups (under 1 per 10,000 through age 29), and increased sharply in older age groups (figure 1). Among men aged 80 years or older, the IFR estimate (95% confidence interval) was 11.6% (8.1% to 16.5%) for confirmed COVID-19 deaths and 16.4% (11.4% to 23.2%) for excess deaths. Among women aged 80 years or older, the corresponding estimates were 4.6% (3.4% to 6.3%) and 6.5% (4.7% to 8.8%).

**Figure 1.**
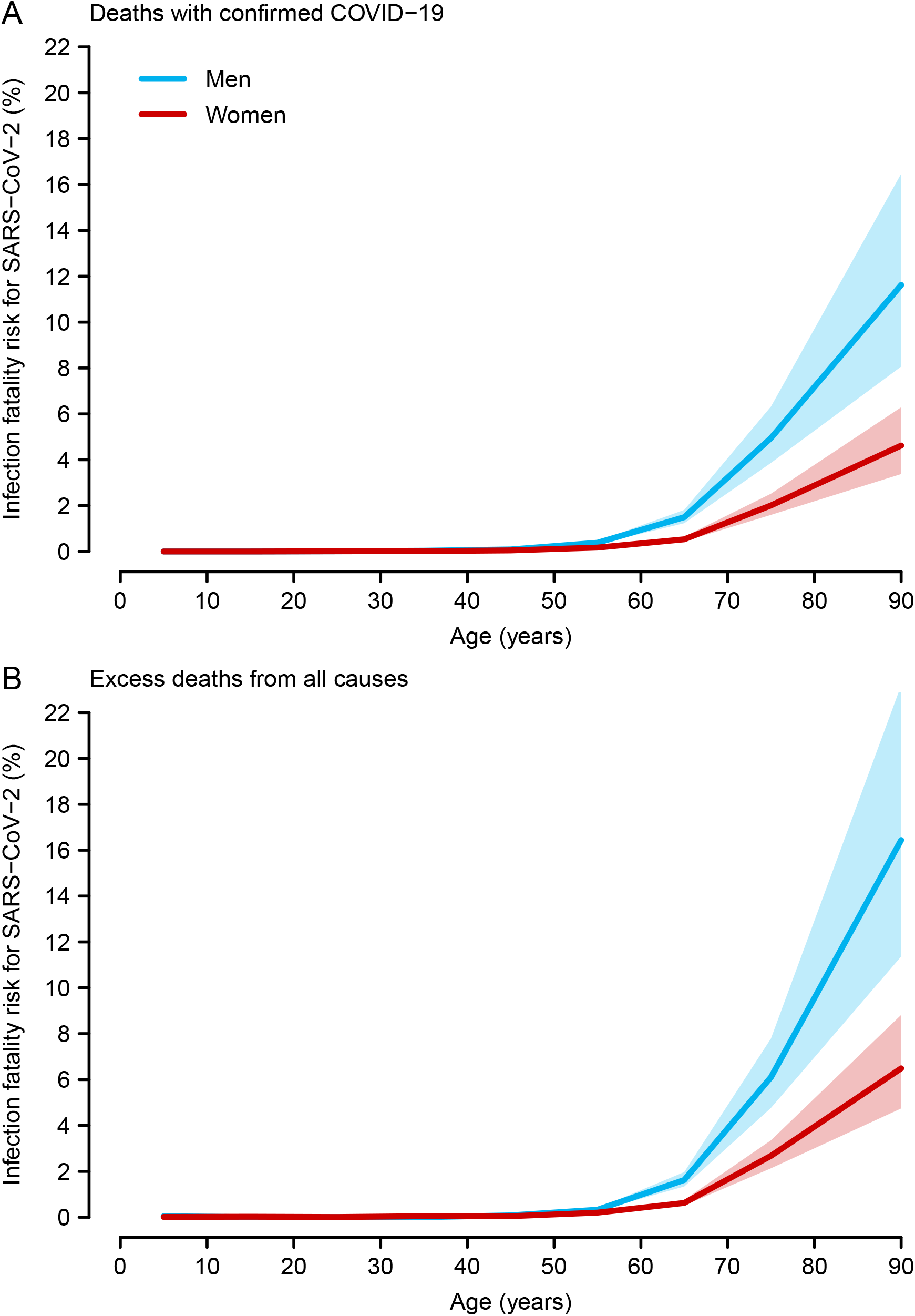
Infection fatality risk for SARS-CoV-2 based on (A) confirmed COVID-19 deaths and (B) excess deaths from all causes in non-institutionalized population, ENE-COVID study, April 27–June 22, 2020, Spain. Shaded regions represent 95% confidence intervals for infection fatality risk.

In sensitivity analyses, the IFR estimates corrected for imperfect sensitivity and specificity were slightly higher, with a corrected overall IFR of 0.88% (95% confidence interval 0.80% to 0.97%) for confirmed COVID-19 deaths and 1.14% (1.03% to 1.25%) for excess deaths (supplementary table 2).

## DISCUSSION

### Principal findings

We estimated an IFR for SARS-CoV-2 between 0.83% and 1.07% in Spain through July 15, 2020. The IFR was greater in men than in women and increased with age: 11.6% to 16.4% in men aged ≥80 years and 4.6% to 6.5% in women aged ≥80 years. Because incomplete ascertainment of deaths is unavoidable during a large-scale epidemic, we obtained separate IFR estimates based on confirmed COVID-19 deaths and excess all-cause deaths. The latter include mortality directly due to SARS-CoV-2 infection and net mortality due to the societal impact of the epidemic and its control measures, such as delayed care for emergencies^14^ and pre-existing chronic conditions due to medical care reorganization and to patients’ reluctance to seek attention,^15,16^ and reductions in traffic injuries and other accidents.^17^

The greater mortality in the elderly may result from a greater prevalence of comorbidities (cardiovascular disease, type 2 diabetes, lung and chronic kidney diseases) that are associated with greater COVID-19 mortality,^18^ and immunological changes (including a decrease of CD8 T cells^19^) that affect the severity of SARS-CoV-2 infections.^20,21^ In addition to greater comorbidity and risk factor prevalence (e.g., smoking), sex differences in cellular immunity may also explain the higher mortality among men, who present a poorer T-cell activation and an increase in pro-inflammatory cytokines.^22^ A negative correlation of T cell response with patients’ age has been reported in males but not in female patients.^22^

### Comparison with previous studies

As mentioned in the introduction, it is difficult to compare IFR estimates from studies using different methods^4^. Case fatality rates –number of deaths divided by the number of confirmed cases-provide different information, as they are heavily influenced by partial case ascertainment and not directly comparable. There are interesting approaches based on modelling,^5^ but models heavily depend on their own assumptions and are not exempt from bias^4^. Our overall IFR estimate is similar to that obtained from serosurveys with low risk of bias^4^.

Our results suggest that some of the heterogeneity in published IFR estimates is driven by the different sex and age structure of the population. Our crude IFR estimates, like others from countries with a similar age-structure, such as Italy,^9,23^ are greater than those from countries with a younger population.^4^ Variations in IFR values might also be related to the local dynamics of the epidemic (e.g., surge in number of new cases, diffusion of the virus among vulnerable collectives), combined with the health system capacity to cope and treat a large number of cases.

### Strengths and limitations

We used data from a nation-wide population-based seroepidemiological study and two complementary sources of mortality information—deaths among laboratory-confirmed COVID-19 cases and excess deaths—to estimate the range of IFR, both overall and by age and sex. The ENE-COVID serosurvey was timed to provide an IFR estimate for first wave of SARS-CoV-2 infection in Spain.^11^ The first round of the study started one month after the peak, which took place around March 20, and the last round ended on June 22. Thus, most participants would have been infected one month before their first participation. As IgG antibodies are detected 2–3 weeks after symptom onset in more than 90% of COVID-19 cases^24^ and decrease 2–3 months after infection,^25^ ENE-COVID is expected to cover infections through at least the first week of June. To include potentially delayed COVID-19 deaths, we considered all deaths registered through July 15 (see supplementary figure 2 for the time distribution of COVID-19 cases and deaths in Spain). Also, the test selected to measure antibodies against SARS-CoV-2 had high sensitivity and very high specificity, according to our meta-analysis (see supplementary figure 1). We used its pooled estimates to calculate corrected ENE-COVID seroprevalence figures; the resulting IFR estimates were slightly higher, but consistent with our primary results (see supplementary methods), showing the robustness of our estimators.

A limitation of the study is that the RENAVE and MoMo surveillance systems did not differentiate deaths from the institutionalized and non-institutionalized population. Hence, we had to collect deaths with confirmed and suspected COVID-19 in nursing homes reported from different Spanish Regional Authorities (supplementary table 1), disaggregate these deaths by sex and age group (as described in supplementary methods), and subtract them from those identified by RENAVE and MoMo. In addition, there is unavoidable uncertainty due to the use of different estimates, but our analyses have incorporated this variability in confidence intervals of the final IFRs (see supplementary methods).

Because the ENE-COVID serosurvey was conducted among the non-institutionalized Spanish population, our IFR estimates do not apply to people living in nursing homes in Spain (about 334,000 residents; 76% being aged 80 or older^26^), where more than 19,000 deaths occurred.^27^ Further research is needed to characterize the mortality in long-term care facilities, which have clusters of vulnerable populations in which the virus may spread very rapidly.^28^ As nursing homes had limited access to hospital care during the initial outbreak in Spain,^29^ our IFR estimates cannot be transported to the institutionalized elderly. Estimating the IFR for SARS-CoV-2 in long-term care facilities will require its own specific approach.^30,31^

### Conclusions

We estimated IFR estimates for SARS-CoV-2 by age and sex in one of the largest serosurveys in the world carried out during the initial outbreak. Our overall IFR estimates (from 0.83% to 1.07%) are about 10 times larger than those for seasonal^32^ or pandemic influenza^33^, in spite of the difficulty of comparing two diseases with mortality figures obtained by different methods^1^. These IFR, considered together with the transmissibility of the disease and the high proportion of susceptible population, provide support for strong control measures.

## Data Availability

The manuscript includes all figures needed to replicate the IFR estimations and indicate the sources used. ENE-COVID seroprevalence figures are provided here for all sex and age groups. Data on deaths come from RENAVE and MoMo, two Spanish National Surveillance Systems. Anonymized data from these systems are available under request. The specific formulary for this purpose is provided by the Department of Communicable Diseases at the National Center for Epidemiology. Instituto de Salud Carlos III. C/ Monforte de Lemos 5 28029 Madrid. ( respectively). Population figures have been provided by the National Institute of Statistics and are publicly available at their website (www.ine.es).

https://portalcne.isciii.es/enecovid19/

https://momo.isciii.es/public/momo/dashboard/momo_dashboard.html

https://cnecovid.isciii.es/covid19/

## ETHICS

ENE-COVID study was approved by the Institutional Review Board of the Institute of Health Carlos III (Register number: PI 39_2020), and a written informed consent was obtained from all participants

## CONTRIBUTORS

RPB, BPG, MAH & MP are responsible for the conception and design of the study; RY & FB are the executive coordinators of the ENE-COVID study; MPO, JOI & AFG are responsible for the serological analysis of the ENE-COVID study, coordinating microbiological labs. JLS, IC, MM, JLP & JFM are responsible for the ENE-COVID study logistics; ILG, PFN, CDS & AL extracted and curated RENAVE and MoMo data; RPB, BPG, MAH, NFL & MP were in charge of statistical analyses and tables and figures design; other authors included in the ENE-COVID group contributed to data acquisition, laboratory analyses and quality control of the ENE-COVID study at their respective regions and/or at national level. The first draft was initially written by RPB, BPG, MAH, MPO, RY, AL & MP. All authors contributed to data interpretation, substantially reviewed the first draft and approved the final version and agreed to be accountable for the work. RPB, BPG & MP act as guarantors, accept full responsibility for the work, had access to the data, and controlled the decision to publish.

## ACKNOWLEDGMENTS

This work was supported by the Spanish Ministry of Health, the Institute of Health Carlos III (Ministry of Science and Innovation) and the National Health System, including the Health Services of all Autonomous Communities and autonomous cities: Servicio Andaluz de Salud, Servicio Aragonés de Salud, Servicio de Salud Principado de Asturias, Servei de Salut Illes Balears, Servicio Canario de la Salud, Servicio Cántabro de Salud, Servicio de Salud Castilla-La Mancha, Servicio de Salud de Castilla y León, Servei Català de Salut, Conselleria de Sanitat Universal i Salut Pública Generalitat Valenciana, Servicio Extremeño de Salud, Servizo Galego de Saúde, Servicio Riojano de Salud, Servicio Madrileño de Salud, Servicio Murciano de Salud, Servicio Navarro de Salud-Osasunbidea & Instituto de Salud Pública y Laboral de Navarra, Servicio Vasco de Salud-Osakidetza and Instituto Gestión Sanitaria. The Spanish Institute of Statistics provided the random selection of households and the information required for participants’ contact. We would like to thank all the nurses, general practitioners, administrative personnel and other health-care workers who collaborated in this study and all participants. This study is the result of the efforts of many professionals and the trust and generosity of more than 60,000 participants who have understood the interest of providing time, information and samples to learn about the situation of the COVID-19 epidemic in our country.

## DATA SHARING

The manuscript includes all figures needed to replicate the IFR estimations and indicate the sources used. ENE-COVID seroprevalence figures are provided here for all sex and age groups. Data on deaths come from RENAVE and MoMo, two Spanish National Surveillance Systems. Anonymized data from these systems are available under request. The specific formulary for this purpose is provided by the Department of Communicable Diseases at the National Center for Epidemiology, Carlos III Institute of Health, Monforte de Lemos 5, 28029 Madrid, Spain (, respectively). Population figures have been provided by the National Institute of Statistics and are publicly available at its website (www.ine.es).

## CODE AVAILABILTY STATEMENT

The code for IFR calculation is available at https://portalcne.isciii.es/ENE-COVID-19/.

## TRANSPARENCY

The lead authors (the manuscript’s guarantors) affirm that the manuscript is an honest, accurate, and transparent account of the study being reported, and no important aspects of the study have been omitted.

